# Risk prediction models for pneumonia in hospitalized stroke patients: A systematic review

**DOI:** 10.1101/2024.11.22.24317773

**Authors:** Manlin Yan, Weirong Huang, Zhihui Zhang, Meixuan Song, Xianrong Li

## Abstract

**Objective:** To systematically evaluate risk prediction models for pneumonia occurrence during hospitalization in stroke patients.

**Methods:** Computer searches were conducted in the PubMed, Embase, Web of Science, Cochrane Library, and EBSCO databases for literature related to risk prediction models for pneumonia in hospitalized stroke patients, with search dates ranging from database inception to June 13, 2024. Two researchers independently screened the literature, extracted the data, and evaluated the risk of bias and applicability of the included studies via the Prediction Model Risk of Bias ASsessment Tool (PROBAST).

**Results:** A total of 43 studies were included, among which 33 studies developed a total of 56 new models, and 25 studies externally validated 19 models. Among the 56 new models, 29 used a logistic regression model (LR), 25 used a machine learning model (ML), 1 used a classification and regression tree model (CART), and 1 used a linear regression model. The reported area under the curve (AUC) ranged from 0.565 to 0.960. The number of predictors explicitly reported for one model was 1,046, with the top three predictors most commonly used being age, the National Institutes of Health Stroke Scale (NIHSS) score, and dysphagia. The PROBAST results revealed that all 43 studies had a high risk of bias, and 27 studies had poor applicability.

**Conclusion:** Although the pneumonia risk prediction models for hospitalized stroke patients in the included studies achieved good predictive performance, the overall quality needs improvement. Future research should follow stricter study designs, standardized reporting practices, and multicenter large-sample external validation.

---

Stroke has become a serious global health issue, ranking as the second leading cause of death worldwide and the third leading cause of death and disability^[1]^. Stroke affects approximately 13.7 million people globally, causing approximately 5.5 million deaths each year^[2]^. Because stroke inpatients have severe conditions, reduced immune and respiratory defense functions, and poor prognoses, up to 30% of stroke inpatients are prone to poststroke infections, with pneumonia being the most common infection^[3, 4]^, which can be categorized as stroke-associated pneumonia (SAP) or ventilator-associated pneumonia. Poststroke pneumonia not only prolongs hospital stays and increases the financial burden on patients’ families but also further aggravates the disease and increases the risk of death for patients. A study including 14,293 stroke patients revealed that the relative risk of death due to poststroke pneumonia is 3.0^[5]^. The development of a pneumonia risk prediction model for hospitalized stroke patients is beneficial for health care workers to identify high-risk individuals early and implement preventive measures, thereby reducing the incidence of pneumonia among stroke patients. Currently, several studies have developed such prediction models, but their predictive performance and overall quality vary. Further validation is needed to assess their clinical applicability. Furthermore, owing to the lack of systematic evaluation of pneumonia risk prediction models for hospitalized stroke patients, the application of high-quality evidence-based medicine is limited. Therefore, we systematically searched and analyzed the relevant literature on pneumonia risk prediction models for hospitalized stroke patients, aiming to provide scientific evidence for the development, application, optimization, and personalized prevention and treatment of poststroke pneumonia in the future.

This study has been registered on the PROSPERO platform (registration number: CRD42024557897).

## 1 Materials and methods

### 1.1 Inclusion and exclusion criteria

#### 1.1.1 Inclusion criteria

The inclusion criteria were as follows: ① the research subjects were stroke patients aged ≥ 18 years; ② pneumonia occurred during hospitalization for stroke; ③ the research content was a construction or validation study of the risk prediction model for pneumonia in hospitalized stroke patients; and ④ the research types were observational studies, such as cohort studies and case-control studies.

### 1.1.2 Exclusion criteria

The exclusion criteria were as follows: ① inability to obtain the full text or incomplete data information ; ② prediction models constructed on the basis of systematic reviews; ③ conference abstracts, academic papers and other informally published literature; ④predictive factors of the model less than 2; ⑤ duplicated published literature; and ⑥ non-English literature.

### 1.2 Search strategy

The computer retrieved the literature related to the risk prediction model of pneumonia in hospitalized stroke patients in the PubMed, Embase, Web of Science, Cochrane Library and EBSCO databases. The retrieval time ranged from the establishment of the database to June 13, 2024. A systematic search was conducted by combining subject terms and free terms, and the search terms used were as follows: stroke, strokes, cerebrovascular accident, CVA, cerebrovascular apoplexy, brain vascular accident, cerebrovascular stroke, apoplexy, cerebral stroke, acute stroke, acute cerebrovascular accident, cerebral infarction, cerebral hemorrhage, brain ischemia, pneumonia, pulmonary infection, pulmonary inflammation, health care-associated pneumonia, stroke-associated pneumonia, risk assessment, risk prediction, prediction, prognosis, forecast, model, tool, score, nomogram model, and nomograph. Taking PubMed as an example, the search strategy is shown in Figure 1.

**Figure.**
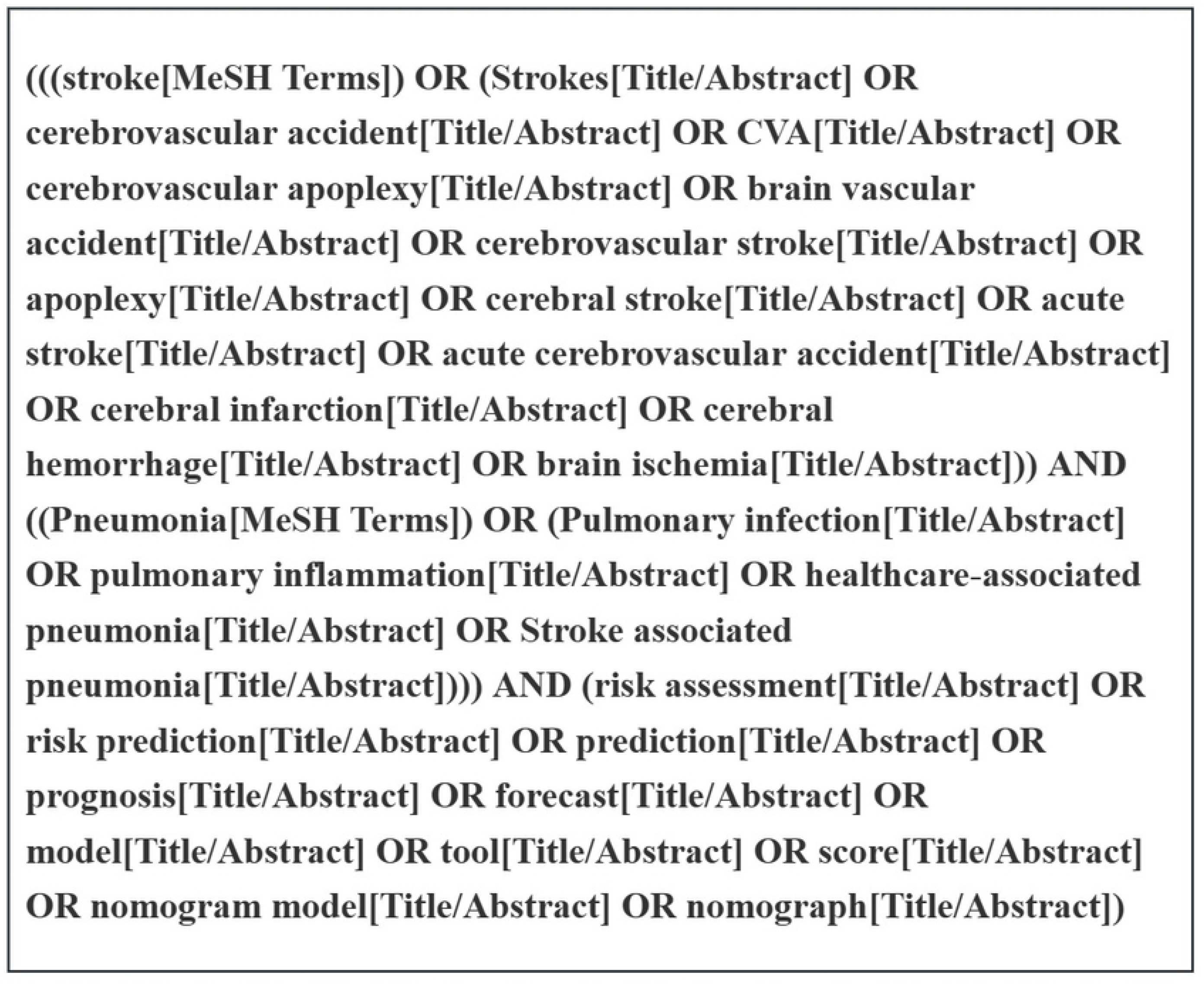

### 1.3 Literature screening and data extraction

After removing duplicate studies via EndNote software, two researchers (MLY and WRH) independently screened the studies and extracted the data in strict accordance with the inclusion and exclusion criteria. The extracted data included the first author, publication time, country, research subjects, research design, modeling method, total sample size, outcome events and incidence rate, area under the receiver operating characteristic curve (AUC), validation method, calibration method, final predictor, etc. In the case of disagreement, the third researcher (XRL) was consulted to reach a consensus.

### 1.4 Assessment of bias risk and applicability of the included studies

Two researchers (MLY and WRH) independently evaluated the bias risks and applicability of the included studies via the Prediction Model Risk of Bias ASsessment Tool (PROBAST)^[6]^ and cross-checked the results. Disagreements were discussed and resolved with a third researcher (XRL).

#### 1.4.1 Assessment of bias risk

The bias risk assessment includes four domains—participants, predictors, outcomes, and analysis—with a total of 20 questions. The answer options are “Yes”, “Probably Yes”, “No”, “Probably No”, and “No Information”. If the answers to all the questions in a certain domain are “Yes” or “Probably Yes”, the bias risk in this domain is low; if the answer to any question is “No” or “Probably No”, the bias risk in this domain is high; if the answer to any question is “No Information” and the answers to other questions are “Yes” or “Probably Yes”, the bias risk in this domain is unclear. If the bias risk in all four domains is low, the overall bias risk is low; if the bias risk in any one domain is high, the overall bias risk is high; if the bias risk in any one domain is unclear and the bias risk in other domains is low, the overall bias risk is unclear.

#### 1.4.2 Application assessment

The applicability assessment includes three domains: participants, predictors, and outcomes. Each domain is evaluated as “low applicability risk”, “high applicability risk”, or “unclear”. If each domain has a “low applicability risk”, the overall applicability risk is low; if any domain has a “high applicability risk”, the overall applicability risk is high; if any domain is “unclear” and the other domains have a “low applicability risk”, the overall applicability risk is unclear.

## 2 Results

### 2.1 Literature search results

After the initial retrieval, 1366 relevant articles were identified. After removing duplicates, 1114 articles remained. Following the review of titles, abstracts, and full texts, a total of 43 articles^[7-49]^ were ultimately included. The literature search flowchart is shown in Figure 2.

**Figure.**
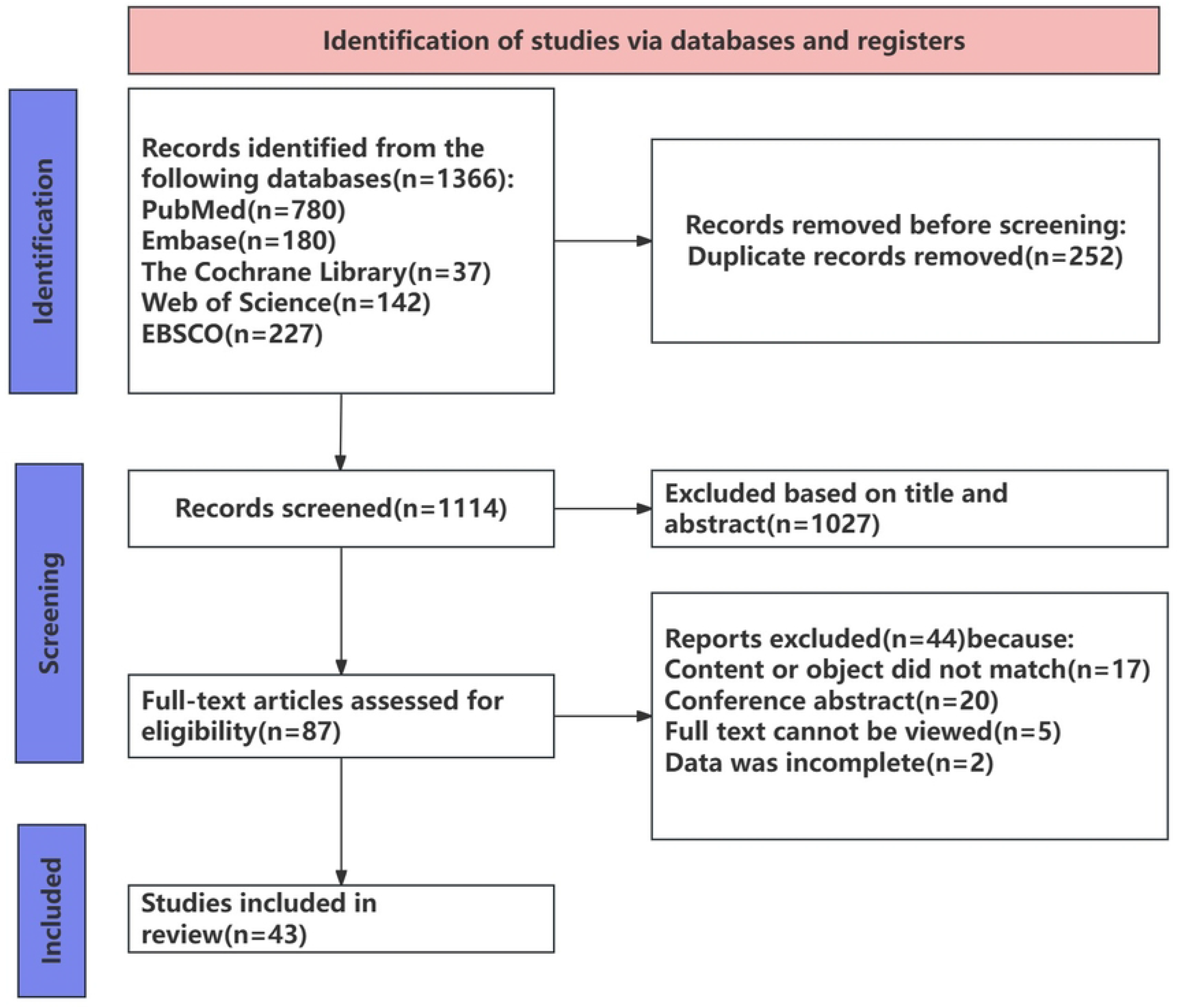

### 2.2 Basic characteristics of the included studies

Among the 43 included studies, most studies were conducted in China (25 studies^[11-13, 17, 22, 25, 26, 28-33, 35, 37-40, 42-46, 48, 49]^); 25 studies^[8, 10, 14, 17, 19-21, 25, 28, 30, 31, 34, 35, 37, 39-49]^were retrospective cohort studies, 16^[7, 9, 11-13, 15, 16, 18, 22-24, 26, 32, 33, 36, 38]^were prospective cohort studies, and 2^[27, 29]^ were retrospective case-control studies. The basic characteristics of the included studies are shown in Table 1.

### 2.3 Basic characteristics of the model

Among the 43 included studies, 33^[7-12, 14, 16, 20, 23, 25-31, 33, 35-49]^ constructed a total of 56 new models. Among them, 29 models used the logistic regression model (LR), 25 models used the machine learning model (ML), 1 model used the classification and regression tree model (CART), and 1 model used the linear regression model. A summary of the studies that constructed new models is shown in Table 2. Among the 43 included studies, 25 studies^[9, 11, 13, 15-19, 21, 22, 24, 26, 28, 30-32, 34, 35, 37-40, 44, 47, 48]^ externally validated the models. Among them, 2 studies^[9, 40]^ included only external validations of the new models they constructed themselves, 2 studies^[11, 38]^ externally validated both the new models they constructed and other known models, and the remaining 21 studies^[13, 15-19, 21, 22, 24, 26, 28, 30-32, 34, 35, 37, 39, 44, 47, 48]^ included only external validations of other known models. A total of 19 models were externally validated. A summary of the studies that conducted external validation of the models is shown in Table 3. The reported AUC ranged from 0.565 to 0.960, indicating that the discrimination ability of the models was good. The number of predictors explicitly reported for one model was 3 to 1,046, and the 3 most commonly used predictors were age, the National Institutes of Health Stroke Scale (NIHSS) score, and dysphagia. A summary of the predictors of the models is shown in Table 4.

### 2.4 Evaluation of bias risks and applicability of the included studies

The PROBAST results revealed that all 43 studies had a high risk of bias, mainly in the domains of research subjects and analysis. For the domain of research subjects, 39 studies^[7-17, 19-23, 25, 27-32, 34-49]^ were rated as high risk. The main reasons were that retrospective studies might have a large recall bias and that the inclusion and exclusion criteria of some studies might be unclear, leading to bias; for the analysis domain, 38 studies^[7-20, 22, 23, 26-31, 33-37, 39-49]^ were rated as high risk. The main reasons might be insufficient sample size, direct exclusion of included subjects with missing data, screening predictors on the basis of univariate analysis methods, failure to assess model discrimination or calibration (or only using the Hosmer–Lemeshow test for calibration), and failure to conduct internal validation (or internal validation only included random split validation of data). The applicability of 27 studies^[7-10, 14-16, 18, 19, 22-24, 27-39, 41, 47]^ was poor. In the domain of research subjects, 23 studies^[7-10, 14-16, 22-24, 27, 29, 30, 32-39, 41, 47]^ were rated as having a high applicability risk. The main reason might be that the inclusion criteria of the participants in the original studies were not clear.

The PROBAST results are shown in Table 5.

## 3 Discussion

### 3.1 The performance of the prediction models is good, but more external validations are needed

The AUC range reported in the 43 included studies was 0.565-0.960, indicating that the discrimination of the models was good and that these models could effectively identify hospitalized stroke patients with pneumonia at high risk. Twenty-six studies^[9-13, 15-17, 20-24, 26, 32, 35, 37, 39, 41-46, 48, 49]^ calibrated the model. The Hosmer-Lemeshow test (H-L) was the most commonly used test (17 studies^[11-13, 15, 17, 21-24, 32, 37, 39, 41, 44, 45, 48, 49]^). The explicitly reported *P* values of H-L were all greater than 0.05, indicating that the predicted results of the models were in good agreement with the actual results. Although these models have shown good performance in validations, the applicability and stability of the models still need to be considered when they are applied in practice.

Among the 43 included studies, 33^[7-12, 14, 16, 20, 23, 25-31, 33, 35-49]^ constructed a total of 56 new models. However, only 33.9% (19/56) of the models were externally validated, and 29 models used the traditional LR method for modeling, making it difficult to avoid correlations among variables, and there was a risk of bias. Furthermore, the study population was predominantly Asian, which might affect the predictive effect of the models in other populations, thereby limiting the universality and extrapolation of the models. Appropriate adjustments might be needed when these models are applied in different regions.

In future research, 25 models in this study should use the ML method for modeling. Some studies have shown that, compared with the traditional LR method, the ML method often yields higher accuracy^[50]^. In this study, the AUC of the models constructed with the ML method ranged from 0.730-0.960, indicating good discrimination and high accuracy of the models. Many studies on risk prediction models face challenges regarding sample size, the processing of continuous variables, and the selection of predictors, some of which can be addressed by integrating the ML method in model development^[51]^. However, a significant drawback of ML at present is the lack of appropriate and simple-to-operate model presentation tools, which limits the clinical application of the models.

In conclusion, the overall performance of the prediction models included in this study is good. However, in future studies, multicenter and large-sample external validations should be conducted to promote the clinical translation and application of the models.

### 3.2 The accuracy and applicability of the models remain to be improved

The PROBAST results indicated that all 43 studies had a high risk of bias, and the applicability of 27 studies^[7-10, 14-16, 18, 19, 22-24, 27-39, 41, 47]^ was poor. The main reasons might be as follows: the majority of the studies were retrospective analyses, which might have large recall bias; the inclusion and exclusion criteria of some studies were not clear; and problems such as insufficient sample size, improper handling of missing values, screening predictors based on univariate analysis methods, failure to conduct standardized performance evaluation and validation of the models, and incomplete reporting.

Therefore, in future studies, the sample size should be expanded, and prospective studies should be adopted as much as possible to ensure the authenticity of the data. When there are missing values, multiple imputation and single imputation methods should be used to reduce the adverse effects of missing data on statistical analysis and model stability^[52]^. Additionally, methods such as LASSO regression and stepwise regression can be adopted for the screening of predictors to ensure that potentially important predictors are not excluded and to improve the predictive performance of the model. Multiple methods or evaluation indicators are used to evaluate the performance of the model to ensure its accuracy. The reporting specifications of PROBAST are used to make the report more transparent and the data more complete. Stricter research methods and data processing should be adopted to improve the accuracy and applicability of the model, thereby providing more reliable risk prediction models for clinical practice.

### 3.3 The collection of predictors is difficult, and there are few common predictors for the models

Among the 56 new models, the number of predictors explicitly reported for one model was 3 to 1,046. For some models, the number of predictors was too large and difficult to collect, resulting in limited clinical applicability of the models. The predictors included in the models of this study mainly consisted of demographic factors, comorbidities, biomarkers and imaging indicators, which have limited potential for clinical intervention. Moreover, obtaining biomarkers and imaging indicators requires professional equipment and technical support, increasing the complexity of the models in clinical application. Therefore, future studies should use more precise algorithms to construct risk prediction models to ensure their accuracy and applicability.

There are few common predictors included in the models of this study. The top 3 most commonly used predictors are age, the National Institutes of Health Stroke Scale (NIHSS) score, and dysphagia. Age is an important risk factor for pneumonia in hospitalized stroke patients. As age increases, the immune function and lung defense ability of patients decline, increasing their susceptibility to infection. Studies have shown that there is a significant correlation between increased age and the occurrence of poststroke pneumonia^[53, 54]^. The NIHSS score is an index used to assess the severity of stroke. The higher the score is, the more severe the brain injury. Studies have shown that hospitalized stroke patients with a high NIHSS score have a greater risk of developing poststroke pneumonia^[54, 55]^, possibly because they have ①more obvious immunosuppression, leading to increased susceptibility to infection, and ② more severe limb paralysis, resulting in an extended bed rest time and an increased chance of aspiration pneumonia, leading to a higher incidence of poststroke pneumonia^[42]^. Studies have shown that the risk of pneumonia in acute stroke patients with dysphagia is 4.08 times greater than that in acute stroke patients without dysphagia^[56]^. Similarly, ^[57]^through a systematic review, Eltringham et al. reported that stroke patients with dysphagia have a greater risk of poststroke pneumonia than do those without dysphagia. The possible reason is that patients with poststroke dysphagia are prone to aspiration of oropharyngeal or gastric contents, thereby causing pneumonia^[3]^. Therefore, early dysphagia screening may help reduce its incidence.

The limitation of this study is that, owing to the large number of included studies and strong heterogeneity, no quantitative analysis was conducted, which may lead to bias in the results of the systematic review. Subsequent studies can include quantitative analyses of risk prediction models for pneumonia in hospitalized stroke patients.

## 4 Conclusion

This study included a total of 43 studies, including 56 models and a systematic evaluation of the characteristics of the models in all aspects. The results suggested that the predictive performance of the risk prediction models for pneumonia in hospitalized stroke patients was relatively good, but the overall risk of bias was relatively high. Future studies should follow more rigorous research designs, include more standardized research reports, and conduct multicenter and large-sample external validations to promote the clinical translation and application of the models. Additionally, it is suggested that clinical medical workers should closely monitor the occurrence of pneumonia in hospitalized stroke patients who are older, have a higher NIHSS score, and have dysphagia, and take timely intervention measures to improve their prognosis.

## Data Availability

All relevant data are within the manuscript and its Supporting Information files.

